# Development and Validation of Nomograms for predicting Coronary Artery Calcification and Severe Coronary Artery Calcification: a retrospective cross-sectional study

**DOI:** 10.1101/2024.09.12.24313598

**Authors:** Peng Xue, Ling Lin, Peishan Li, Zhengting Deng, Xiaohu Chen, Yanshuang Zhuang

**Affiliations:** Department of Geriatrics Cardiology, Taizhou Affiliated Hospital of Nanjing University of Chinese Medicine, 86 Jichuan East Road, Taizhou, Jiangsu, 225300, People’s Republic of China; Department of Chinese Medicine, Nanjing Brain Hospital, 264 Guangzhou Road, Nanjing, Jiangsu, 210029, People’s Republic of China; Nanjing University of Chinese Medicine, 138 Xianlin Avenue, Nanjing, Jiangsu, 210029, People’s Republic of China; Department of Geriatric Cardiology, Jiangsu Province Hospital of Chinese Medicine, 155 Hanzhong Road, Nanjing, Jiangsu, 210029, People’s Republic of China; Department of the science and technology, Taizhou Affiliated Hospital of Nanjing University of Chinese Medicine, 86 Jichuan East Road, Taizhou, Jiangsu, 225300, People’s Republic of China

**Keywords:** Coronary artery calcification, Nomogram, Predictive model, LASSO

## Abstract

**Introduction:** There is a significant lack of effective pharmaceutical interventions for treating coronary artery calcification (CAC). Severe CAC (sCAC) poses a formidable challenge to interventional surgery and exhibits robust associations with adverse cardiovascular outcomes. Therefore, it is imperative to develop tools capable of early-stage detection and risk assessment for both CAC and sCAC. This study aims to develop and validate nomograms for the accurate prediction of CAC and sCAC.

**Methods:** This retrospective cross-sectional study was conducted in Taizhou, Jiangsu Province, China. CAC assessment was performed using non-gated thoracic CT scans. Demographic data and clinical information were collected from patients who were then randomly divided into a training set (70%) or a validation set (30%). Least absolute shrinkage and selection operator (LASSO) regression as well as multiple logistic regression analyses were utilized to identify predictive factors for both CAC and sCAC development. Nomograms were developed to predict the occurrence of CAC or sCAC events. The models’ performance was evaluated through discrimination analysis, calibration analysis, as well as assessment of their clinical utility.

**Results:** This study included 666 patients with an average age of 75 years, of whom 56% were male. 391 patients had CAC, with sCAC in 134 cases. Through LASSO and multiple logistic regression analysis, age increase, hypertension, carotid artery calcification, CHD, and CHADS_2_ score were identified for the CAC risk predictive nomogram with an area under the receiver operating characteristic (ROC) curve (AUC) of 0.845(95%CI 0.809-0.881) in the training set and 0.810(95%CI 0.751-0.870) in the validation set. Serum calcium level, carotid artery calcification, and CHD were identified for the sCAC risk predictive nomogram with an AUC of 0.863(95%CI 0.825-0.901) in the training set and 0.817(95%CI 0.744-0.890) in the validation set. Calibration plots indicated that two models exhibited good calibration ability. According to the decision curve analysis (DCA) results, both models have demonstrated a positive net benefit within a wide range of risks.

**Conclusions:** The present study has successfully developed and validated two nomograms to accurately predict CAC and sCAC, both of which have demonstrated robust predictive capabilities.

## INTRODUCTION

Coronary artery calcification (CAC), a highly specific hallmark of coronary atherosclerosis^1^, has been extensively investigated and is widely accessible in the field of cardiovascular medicine. CAC, being the most predictive single cardiovascular risk marker in asymptomatic individuals, can provide supplementary prognostic information in addition to traditional cardiovascular risk factors^2, 3^. Patients with severe coronary artery calcification (sCAC) are less likely to undergo complete revascularization and exhibit a poorer clinical prognosis, thus establishing sCAC as an independent predictor of adverse outcomes^4^.

There is currently no evidence to suggest that medication can improve CAC. Despite the application of surgical interventions such as rotational atherectomy and intracoronary lithotripsy to improve CAC^5–7^, which is advantageous for stent implantation, significant challenges still persist in the prevention and treatment of CAC. Therefore, it is of great significance to screen for coronary artery calcification and develop corresponding preventive strategies. Due to limited resources, it is of great clinical significance to quickly and effectively identify high-risk patients with CAC or sCAC. Clinical prediction models can aid physicians in assessing the risk of CAC and sCAC, thereby providing guidance for conducting screening for CAC.

Several studies have reported risk factors associated with coronary artery calcification, including increasing age, variability in fasting blood glucose levels, smoking, hyperlipidemia, hypertension, and diabetes^8–15^. In addition, chronic kidney disease (CKD) has also been confirmed as a risk factor for CAC and shows a progressively worsening trend with the severity of CKD^16, 17^. A predictive model has been established to accurately predict sCAC, primarily applicable to patients with end-stage renal failure ^18^. Meanwhile, a machine learning model has also been developed to predict CAC. However, this model analyzes a smaller number of variables and may have confounding factors that cannot be excluded, resulting in suboptimal predictive performance and limited clinical utility^19^.

In this retrospective cross-sectional study, we conducted a comprehensive analysis of nearly 30 factors and employed least absolute shrinkage and selection operator (LASSO) regression to select variables, thereby mitigating the impact of collinearity. Our objective was to develop two concise and pragmatic nomograms for predicting CAC and sCAC, to aid clinical practitioners in identifying high-risk populations.

## METHODS

### Study population and design

The retrospective study was conducted at Taizhou Hospital of Traditional Chinese Medicine in Jiangsu Province, China. The study cohort comprised patients admitted to the hospital between September 2022 and July 2024, all of whom underwent chest CT examination during their hospitalization. Exclusion criteria included: 1. History of coronary stent implantation; 2. End-stage renal disease or regular dialysis treatment; 3. Incomplete clinical information. The requirement for patient consent was waived due to the retrospective nature of the study design. We ensured that the data were properly anonymized and maintained confidentiality, adhering to the principles outlined in the Declaration of Helsinki.

### Definitions of CAC and sCAC

In comparison to electrocardiogram gating, non-gated chest CT scans for coronary artery calcium score (CACS) exhibit a remarkable concordance in predicting cardiovascular events^20, 21^. We employed non-gated chest CT scans to acquire the CACS and utilized the Agatston method for determining CAC (CACS>0) and sCAC (CACS>400)^22^.

### Data collection

The data used in this study were collected from electronic medical records, including demographics (gender, age, economic status, height, weight, smoking and drinking history), laboratory test results (low-density lipoprotein cholesterol level, high-density lipoprotein cholesterol level, triglyceride level, uric acid level, serum creatinine concentration, serum urea nitrogen concentration, serum calcium concentration, calcium-phosphorus product value, serum bicarbonate concentration, serum total bilirubin level, alkaline phosphatase activity index, blood platelet count and hemoglobin A1c), comorbidconditions (insomnia, hypertension, diabetes mellitus, coronary heart disease, chronic heart failure, chronic obstructive pulmonary disease, ischemic cerebrovascular disease, carotid artery calcification plaque), CHADS_2_ score and CACS. If multiple laboratory tests were performed during hospitalization, the first test result within 24 hours after admission was selected.

### Feature selection

Based on clinical experience and literature research, we have preliminarily identified key variables potentially associated with CAC. The variables required for predicting CAC are consistent with those used in sCAC. To address issues of collinearity and overfitting, we employed LASSO regression analysis to screen all variables and select the predictive ones. Restricted cubic spline (RCS) regression was employed to explore potential nonlinear relationships between the nonselected continuous variables and CAC or sCAC.

### Model development and visualisation

The data were randomly divided into a training set and a validation set in a 7:3 ratio to construct the CAC and sCAC models. The training set was utilized for model development, while the validation set served for model validation. Logistic regression analysis was performed using predictive variables selected by LASSO regression to establish these models. The application of logistic regression models was demonstrated through constructing two nomograms. The SHapley Additive exPlanation (SHAP) method was employed to present a ranking of variable importance in two models.

### Model evaluation and validation

Discrimination, calibration, and clinical applicability were utilized for the evaluation of model predictive performance. The receiver operating characteristic (ROC) curve was employed to assess model discrimination. Evaluation of model performance included analysis of the area under the curve (AUC), specificity, and sensitivity. The calibration of the model was evaluated using the Hosmer-Lemeshow (H-L) test and calibration plot. When the p-value of the H-L test is greater than 0.05, it indicates that the model has a strong level of predictive accuracy. Clinical applicability was evaluated using decision curve analysis (DCA) and clinical impact curve (CIC).

### Statistical analyses

The data analysis was performed using R version 4.3.0. Patients were categorized into two groups based on the presence or absence of CAC or sCAC, as presented in Table 1. Normally distributed continuous variables were reported as mean and standard deviation (SD), while non-normally distributed continuous variables were expressed as median and interquartile range (IQR). Categorical variables were represented by patient count and percentage for each variable. A t-test was employed for normally distributed continuous variables, whereas a Mann-Whitney U test was utilized for non-normally distributed ones. Proportion comparison tests were conducted using either the chi-square test with Yates correction or Fisher’s exact test. Statistical significance was defined as a p-value less than 0.05 (two-sided).

**Table 1.**
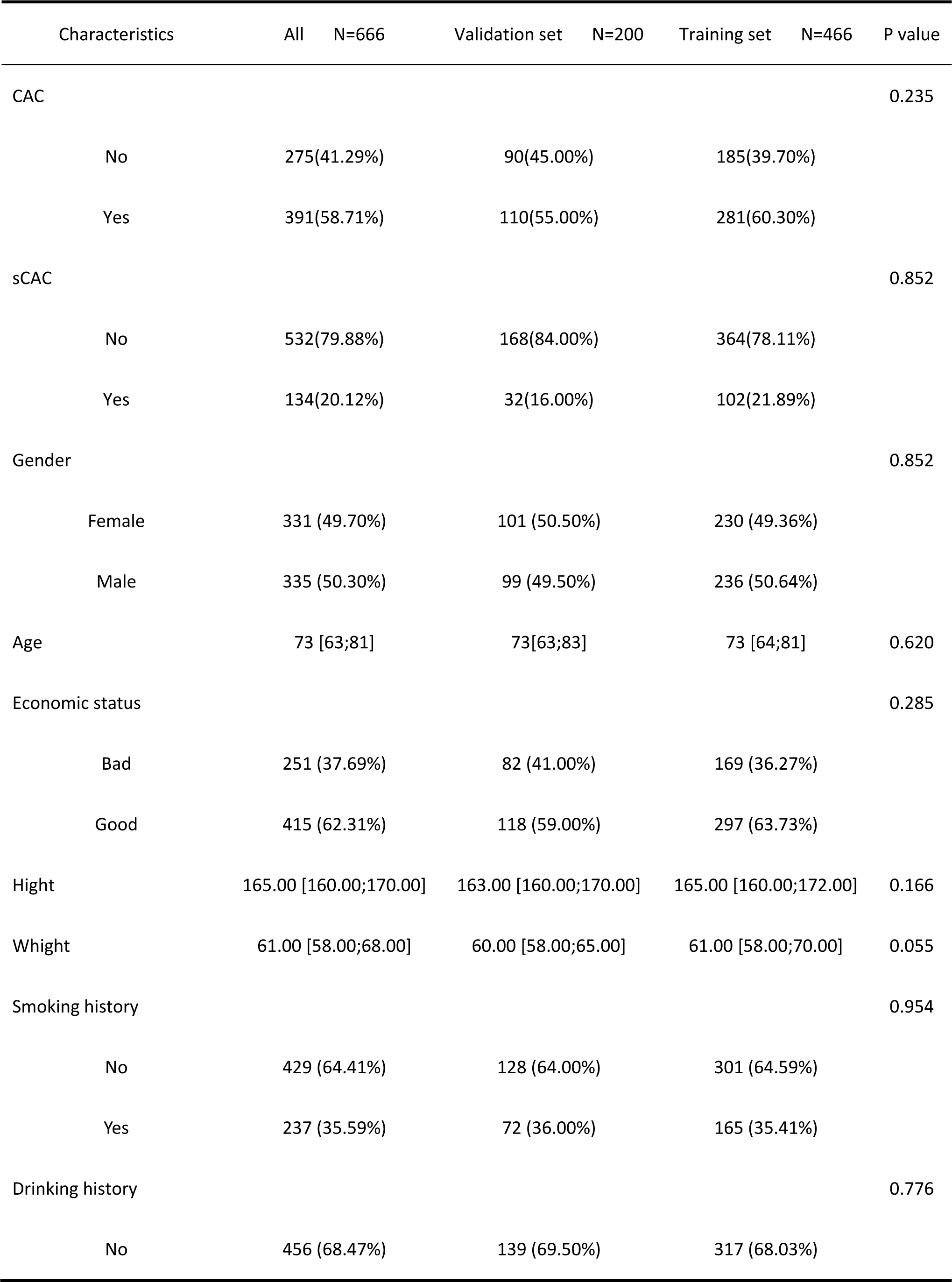
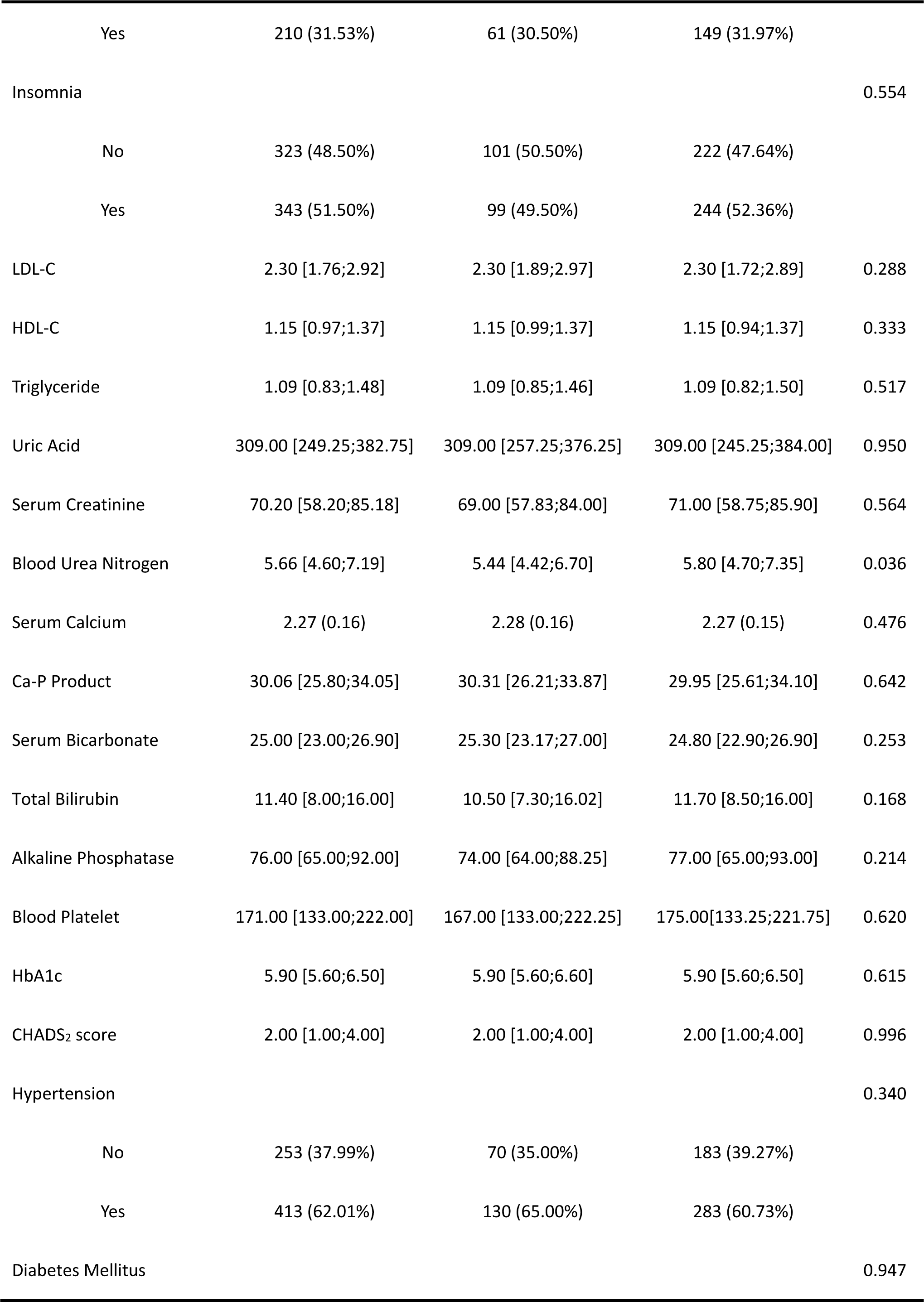
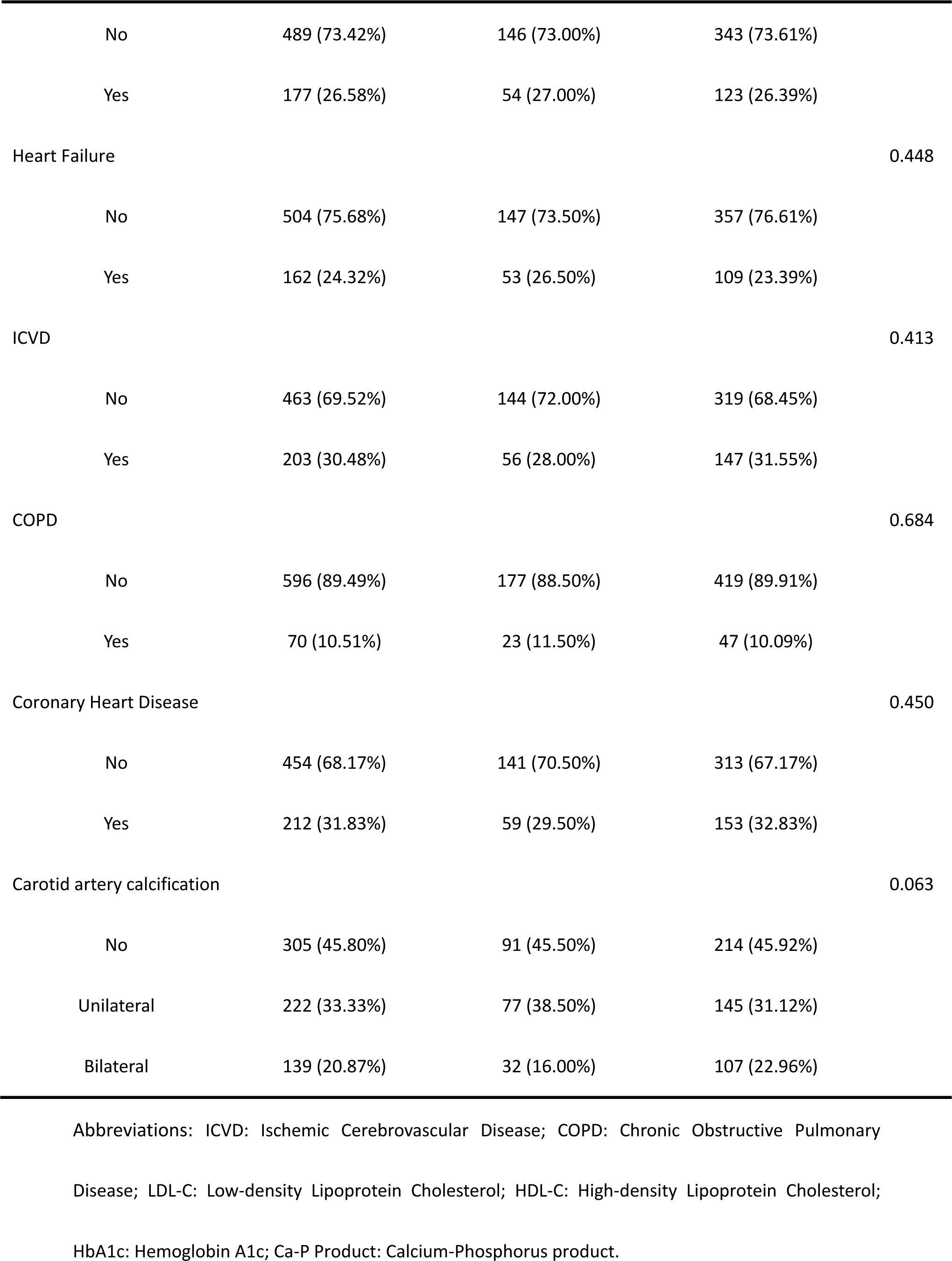
Baseline characteristics of the study subjects.

## RESULTS

### Patient characteristics

A total of 666 patients were included in the study (Figure 1). The average age of these patients was 73 years (interquartile range: 63-81), with males accounting for 50.30% of the group. Among all participants, CAC was observed in 391 individuals (58.71%), while sCAC occurred in 134 cases (20.12%). Table 1 demonstrates a well-balanced distribution of patient characteristics between the training set (466 cases, 70%) and validation set (200 cases, 30%).

**Figure 1.**
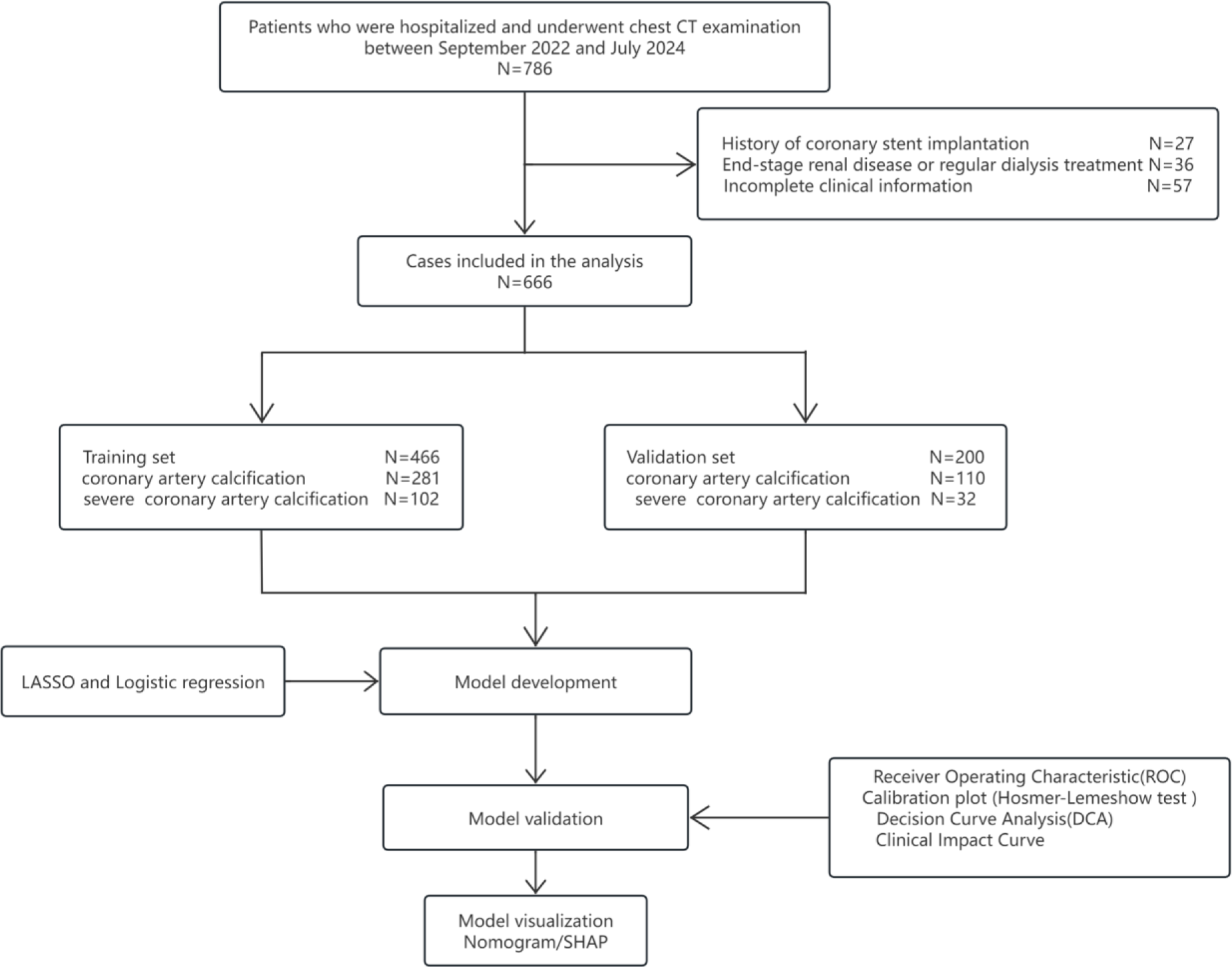
Flowchart of the steps for predicting Coronary Artery Calcification and severe Coronary Artery Calcification.

### Feature selection

The present study investigated a total of 29 variables, utilizing LASSO regression for variable selection and performing 10-fold cross-validation. The findings unveiled significant associations between age, hypertension, carotid artery calcification, coronary heart disease (CHD), and CHADS_2_ score with CAC. Moreover, age, serum calcium level, carotid artery calcification, and CHD were identified as potential predictors for sCAC (Figure 2).

**Figure 2.**
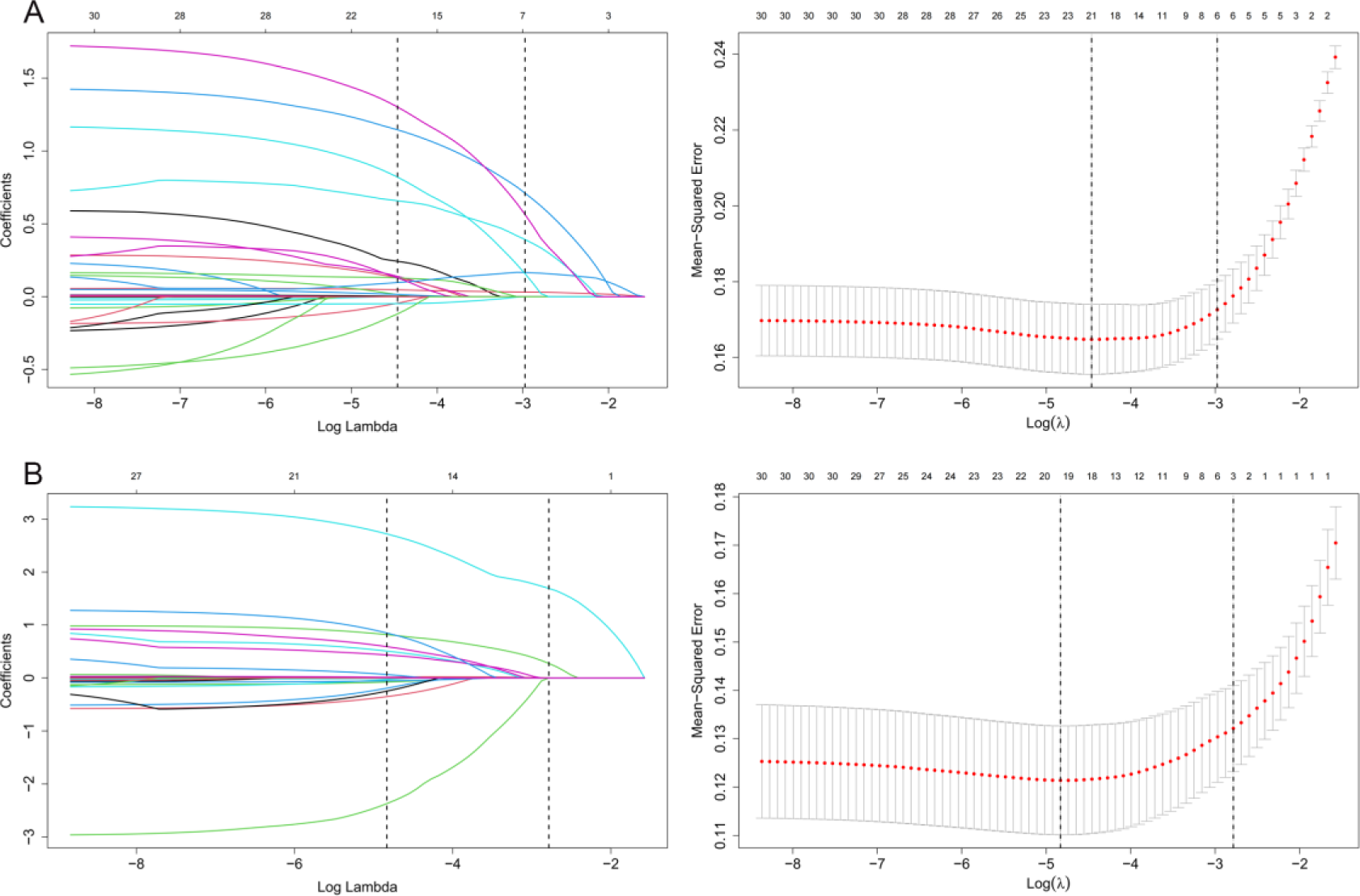
Variable selection is performed using the Least Absolute Shrinkage and Selection Operator (LASSO). The left plots display coefficient profile plots against a log(lambda) sequence, illustrating the variable selection process by identifying nonzero coefficients through optimal lambda derivation. The right plots depict the best matching factors identified through ten-fold cross-validation, with lambda.1se used as the criterion for factor selection. (A) Coronary Artery Calcification; (B) severe Coronary Artery Calcification.

### Model development and visualisation

The variables selected via LASSO regression were utilized to construct predictive models for CAC and sCAC using multiple logistic regression analysis, employing the backward stepwise method. According to the results from Table 2, age (OR=1.041, 95%CI 1.017-1.066), hypertension (OR=2.152, 95%CI 1.236-3.763), unilateral carotid artery calcification (0R=2.515, 95%CI 1.478-4.311), bilateral carotid artery calcification (OR=4.782, 95%CI 2.441-9.845), CHADS_2_ score (OR=1.229, 95%CI 1.013-1.499), and CHD (OR=3.792, 95%CI 6.647-22.26) were identified as independent risk factors for CAC. On the other hand, serum calcium level (0R=0.051, 95%CI 0.008-0.295), CHD (OR=2.851, 95%CI 1.667-4.922), unilateral carotid artery calcification (OR=3.637, 95%CI 1.751-8.023) and bilateral carotid artery calcification (OR=25.40, 95%CI 12.50-55.98) were predictive factors for sCAC.

**Table 2.** Multivariate logistic regression analysis of predictors selected by LASSO regression procedure.

| Model | Characteristics | B | SE | OR | CI | Z | P |
| --- | --- | --- | --- | --- | --- | --- | --- |
| CAC | (Intercept) | -4.332 | 0.80492 | 0.013 | 0.013 (0.002-0.061) | -5.382 | <0.001 |
|  | Age | 0.04 | 0.01198 | 1.041 | 1.041 (1.017-1.066) | 3.363 | 0.001 |
|  | CHD | 1.333 | 0.27819 | 3.793 | 3.792 (2.226-6.647) | 4.792 | <0.001 |
|  | Uni-Carotid Artery Calcification | 0.922 | 0.2725 | 2.515 | 2.515 (1.478-4.311) | 3.385 | 0.001 |
|  | Bil-Carotid Artery calcification | 1.565 | 0.35375 | 4.782 | 4.782 (2.441-9.845) | 4.424 | <0.001 |
|  | CHADS<sub>2</sub> Score | 0.207 | 0.09972 | 1.23 | 1.229 (1.013-1.499) | 2.075 | 0.038 |
|  | Hypertension | 0.767 | 0.28344 | 2.153 | 2.152 (1.236-3.763) | 2.705 | 0.007 |
| sCAC | (Intercept) | 3.406 | 2.01815 | 30.151 | 30.15 (0.583-1622.) | 1.688 | 0.091 |
|  | Serum Calcium | -2.962 | 0.9012 | 0.052 | 0.051 (0.008-0.295) | -3.287 | 0.001 |
|  | CHD | 1.048 | 0.27535 | 2.852 | 2.851 (1.667-4.922) | 3.806 | <0.001 |
|  | Uni-Carotid Artery Calcification | 1.291 | 0.38478 | 3.638 | 3.637 (1.751-8.023) | 3.356 | 0.001 |
|  | Bil-Carotid Artery Calcification | 3.235 | 0.37987 | 25.401 | 25.40 (12.50-55.98) | 8.516 | <0.001 |

Abbreviations : CAC: Coronary Artery Calcification; sCAC: Severe Coronary Artery Calcification; CHD:
Coronary Heart Disease

We developed two nomograms to visually represent these predictive models (Figure 3). Each predictive variable is assigned a corresponding score, and by summing the scores of all variables and mapping it onto a calibrated scale, we can draw a vertical line to determine the probability of predicting CAC and sCAC risk. In the prediction of the CAC model, CHD demonstrates the highest importance, followed by carotid artery calcification, CHADS_2_ score, age, and hypertension. Meanwhile, in the prediction of the sCAC model, carotid artery calcification surpasses both CHD and serum calcium in terms of variable importance.

**Figure 3.**
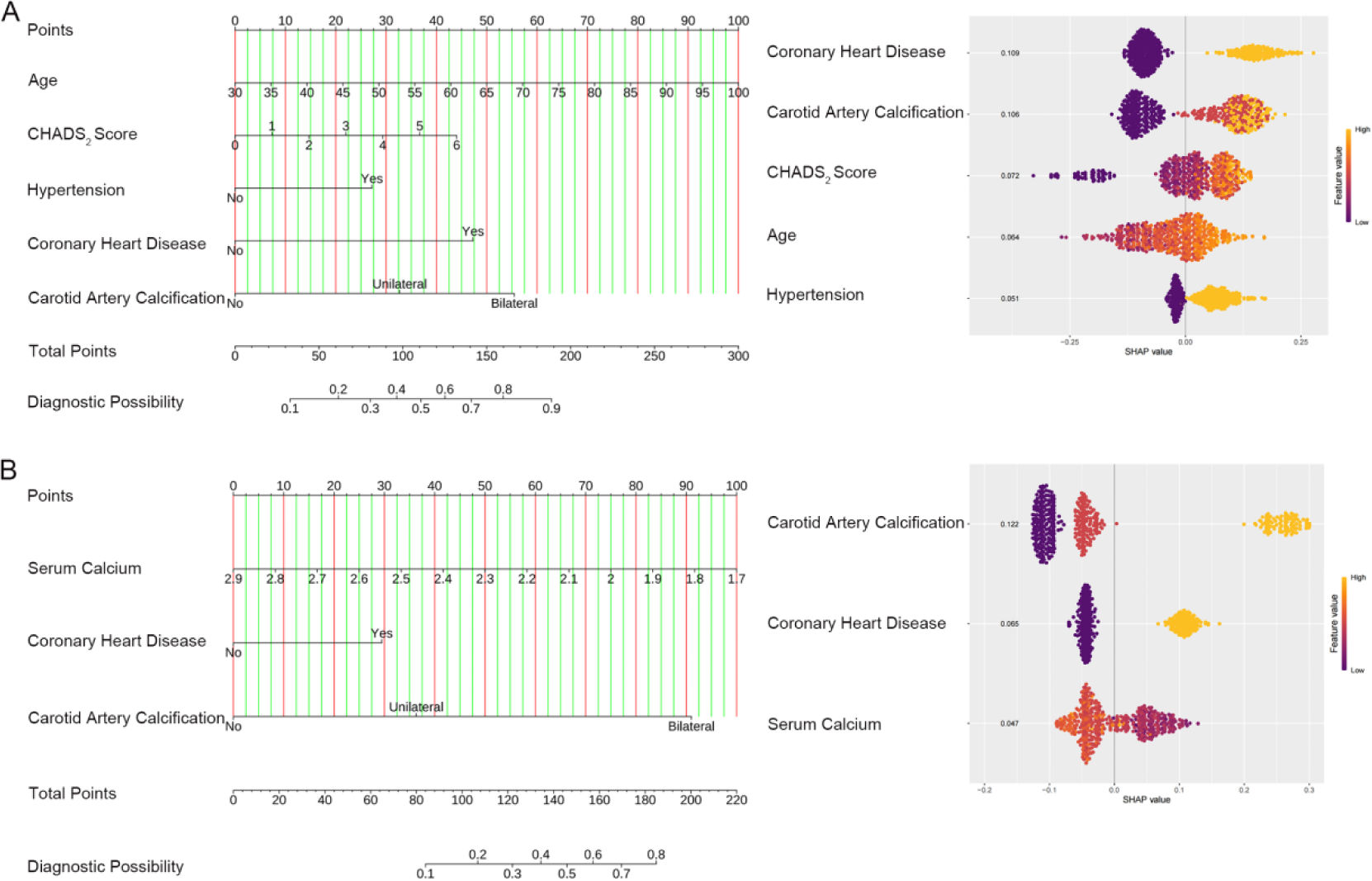
The left plots are nomograms to predict the probability of CAC (A) and sCAC (B). A total score was generated by summing the scores for each predictor. Total score corresponding probability of CAC or sCAC. The right plots illustrate the models’ interpretation using the SHAP method, providing a succinct summary of feature contributions through SHAP values. The density scatter plot visually represents all samples, while the ranking of features is determined based on their cumulative absolute SHAP values. CAC, Coronary Artery Calcification; sCAC, severe Coronary Artery Calcification.

### Model evaluation and validaiton

The ROC curves of the two Nmograms, including cutoff points and specificity, sensitivity, are presented in Figure 4. In the training set, the AUC for predicting CAC was 0.845 (95%CI 0.809-0.881), while in the validation set it was 0.810 (95%CI 0.751-0.870) (Figure 4A). For predicting sCAC, the AUC was found to be 0.863 (95%CI 0.825-0.901) in the training set and 0.817 (95%CI 0.744-089) in the validation set (Figure 4B).

**Figure 4.**
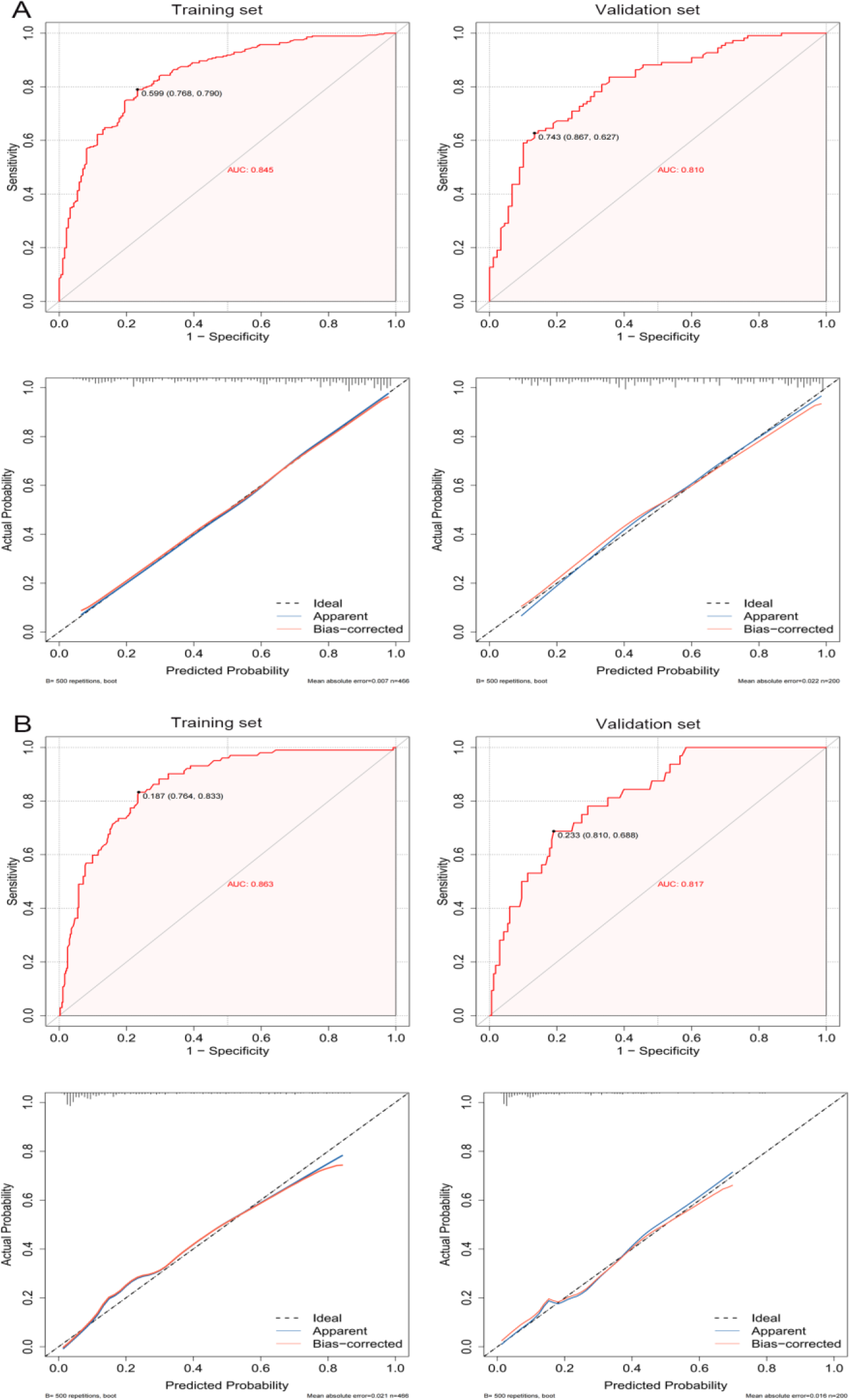
shows the discrimination and calibration of nomograms for predicting CAC (A) and sCAC(B). The ROC curve (upper) display the AUC and the corresponding cut-off points. The calibration plot (lower) represents the actual observed probability on the Y-axis, while the model-predicted probability is shown on the X-axis. CAC, Coronary Artery Calcification; sCAC, severe Coronary Artery Calcification.

The calibration plots based on the Hosmer-Lemeshow (H-L) test are presented in Figure 4. The p-values for predicting CAC using the H-L test were 0.983 and 0.211 for the training set and validation set, respectively; while for predicting sCAC, they were found to be 0.890 and 0.859 in turn for these two sets of data as well. These results indicated that both models demonstrate high accuracy.

The DCA curves demonstrated that both models exhibit positive net benefits across a wide range of risk (Figure 5). In the validation set, the threshold probabilities for predicting CAC and sCAC were 12% to 88% and 2% to 66%, respectively (Figure 5A and 5B). The CIC revealed that when the risk thresholds exceeded 0.55 and 0.35, respectively, both models demonstrated a strong concordance between predictions and actual occurrences in the validation set, indicating significant clinical predictive efficacy (Figure 5A and 5B).

**Figure 5.**
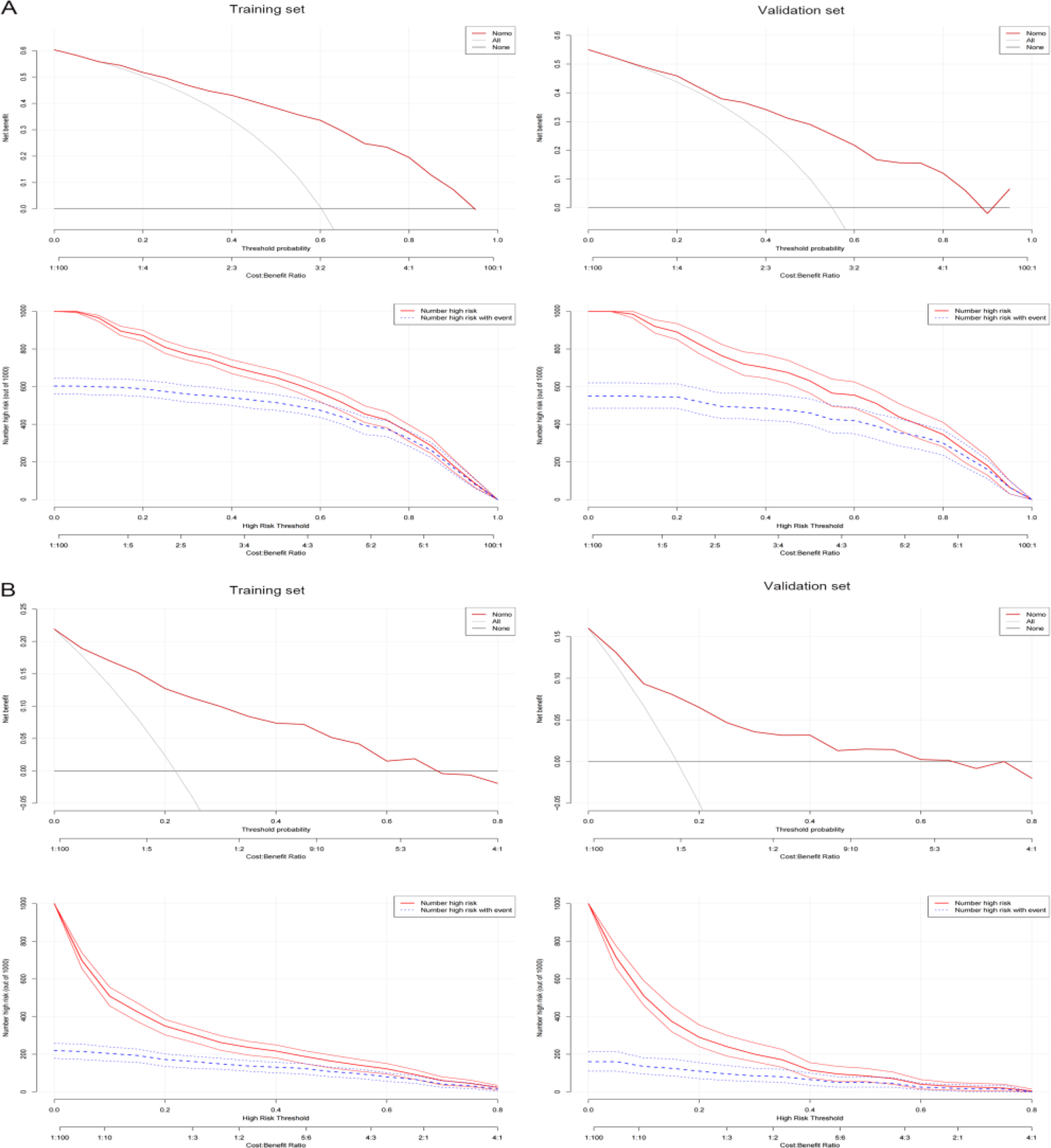
shows the decision curve analysis (DCA) and clinical impact curve (CIC) of nomograms for predicting CAC (A) and sCAC (B). The DCA curve (upper) illustrates the extent of the model’s adaptability. The Y-axis shows the net benefits. The thick solid line indicates that all patients were assumed to be free from CAC or sCAC, and the thin solid line indicates that all patients were assumed to have CAC or sCAC. The red solid line indicates the risk nomogram. The clinical impact curve of the model; The X-axis represents the threshold for high-risk, while the Y-axis represents the number of individuals at risk per 1,000. The red curve depicts the model’s predicted count of individuals experiencing the event, whereas the blue curve illustrates the actual count of individuals who have experienced it. CAC, Coronary Artery Calcification; sCAC, severe Coronary Artery Calcification.

### Correlation between HDL-C and sCAC

The RCS analysis did not identify any continuous variables that exhibited a nonlinear relationship with CAC (Supplementary Material 1); however, we discovered a significant nonlinear association between HDLC and sCAC (Supplementary Material 2 and Figure 6A). This study suggested that HDL-C levels below 0.73mmol/L or above 1.35mmol/L confer protection against sCAC, while HDL-C levels ranging from 0.73mmol/L to 1.35mmol/L were considered as risk factor (Figure 6A). We included HDL-C as a categorical variable in the multivariable model for analysis based on two cutoff points, 0.73mmol/L and 1.35mmol/L. Compared to the model that only included serum calcium, CHD, and carotid artery calcification as predictors of sCAC, there was no statistically significant difference in the AUC (0.866 vs. 0.863, P=0.547) (Figure 6B).

**Figure 6.**
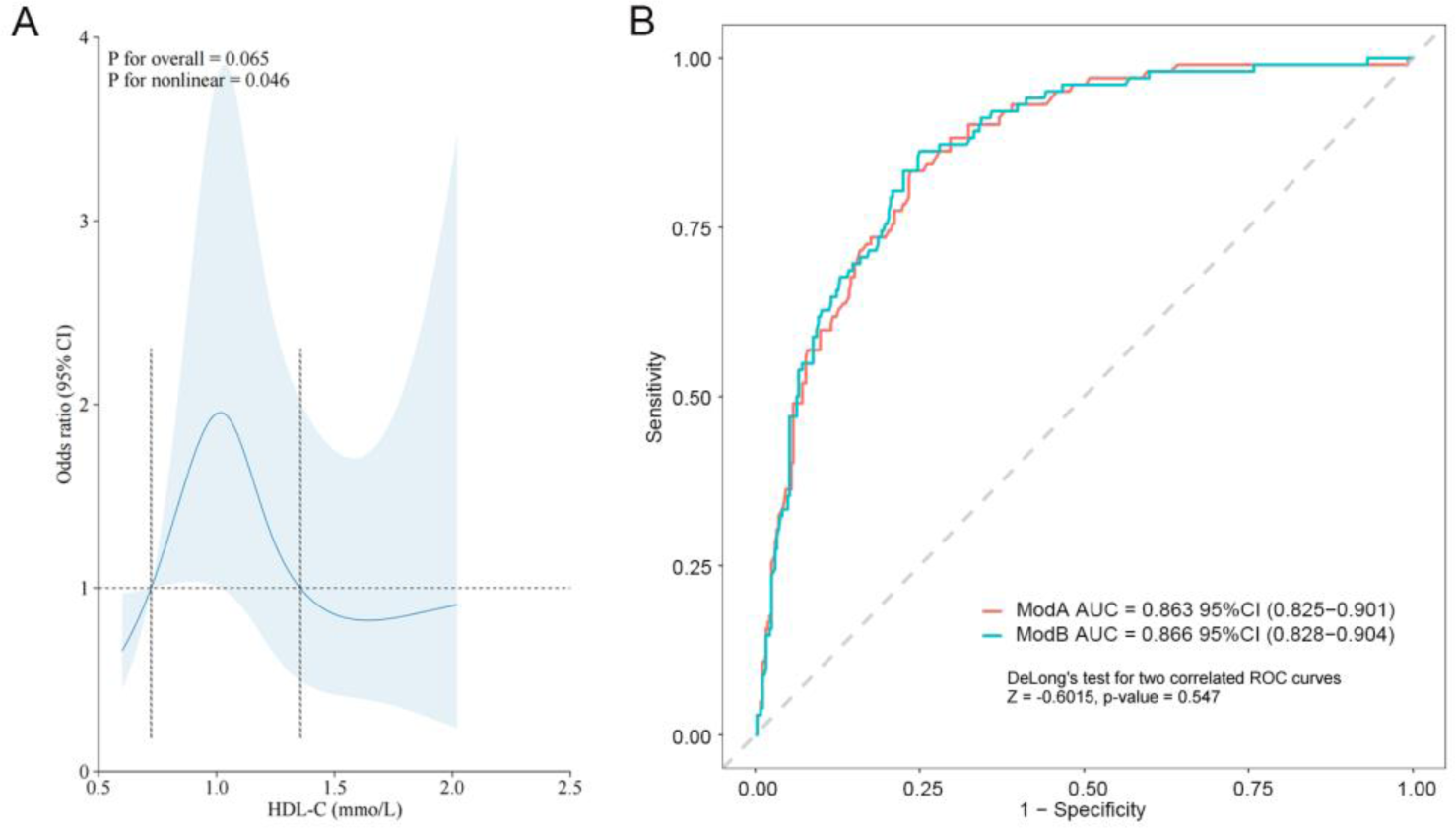
(A) Association Between HDL-C and sCAC Using a Restricted Cubic Spline Regression Model. Graphs show ORs for sCAC according to HDL-C adjusted for serum calcium, coronary heart disease, carotid artery calcification. Data were fitted by a logistic regression model, and the model was conducted with 4 knots at the 5th, 35th, 65th, 95th percentiles of HDL-C (reference is the 5th percentile). Solid lines indicate ORs, and shadow shape indicate 95% CIs. (B) Comparison of models. Model A was constructed based on coronary heart disease, serum calcium, and carotid artery calcification, while model B further incorporates the influence of HDL-C. OR, odds ratio; CI, confidence interval. sCAC, severe Coronary Artery Calcification; HDL-C, High-density Lipoprotein Cholesterol.

## DISCUSSION

In this study, we developed and validated two predictive nomograms to assess the risk of CAC and sCAC in all-cause hospitalized patients. The findings indicate that age, coronary heart disease(CHD), carotid artery calcification, CHADS_2_ score, and hypertension serve as predictors for CAC, while serum calcium level, CHD, and carotid artery calcification are predictors for sCAC. Through evaluation of the models’ discriminative ability, calibration performance, and clinical applicability assessment; both predictive nomograms demonstrate robust predictive capabilities. Importantly, all the predictive indicators can be obtained through non-invasive examinations such as medical history review along with blood tests, and color Doppler ultrasound examination. Ultimately a clinical decision can be made regarding the need for further improvement by performing chest CT or coronary CT angiography examinations.

CAC was associated with an elevated risk of cardiovascular disease, necessitating active screening in both high-risk and low-risk populations for Cardiovascular disease (CVD)^3^. The prevalence and spectrum of CAC among middle-aged and elderly patients were progressively increasing ^23^, thus warranting special attention. This study revealed an independent association between age and the risk of CAC, with a 4.1% increase in risk for every one-year increment in age. However, it is noteworthy that age was not included as a covariate in the severe calcification model, suggesting that its correlation with sCAC remains uncertain. In the population of patients with end-stage renal failure, age was identified as an independent risk factor for sCAC^18, 24^, indicating potential heterogeneity within the study cohort. In recent years, there has been a paucity of research investigating the risk factors associated with sCAC in non-renal disease populations, necessitating further exploration into the relationship between age and sCAC.

Carotid artery calcification was considered a significant risk factor for stroke^25^.A meta-analysis revealed that the presence of carotid plaques with accompanying calcification, even without stenosis, increases the risk of ipsilateral ischemic stroke by 1.72 times^26^. However, there remains ongoing debate regarding whether carotid plaque calcification independently contributes to cardiovascular events^27–29^. Currently, there is a paucity of research investigating the utility of carotid artery calcification as a prognostic indicator for CAC. Our findings demonstrate that bilateral carotid artery calcification confers an increased risk of both CAC and sCAC when compared to unilateral carotid artery calcification. These results suggest a correlation between the degree of carotid artery calcification and the extent and severity of CAC. Consequently, we recommend utilizing color Doppler ultrasound for carotid artery screening to determine the necessity of additional chest CT or coronary artery CTA examinations, thereby evaluating the extent of CAC. Moreover, in kidney transplant recipients, Carotid-femoral pulse wave velocity could potentially serve as an indicator for predicting the presence of severe coronary artery calcification^30^. Therefore, future research should aim to further investigate the association between vascular functional status subsequent to vascular calcification and coronary artery calcification.

The CHADS_2_ scoring system (1 point for chronic heart failure, hypertension, advanced age, and diabetes, and 2 points for previous stroke/transient ischemic attack) was used to predict the risk of cardioembolic stroke in patients with atrial fibrillation. In recent years, studies have demonstrated the utility of the CHADS_2_ score not only in predicting cardiovascular events and all-cause mortality among non-atrial fibrillation patients^31^, but also in forecasting all-cause mortality associated with COVID-19^32^. However, there is currently a dearth of research investigating the application of the CHADS_2_ scoring system for predicting coronary artery calcification. Our research findings suggest a positive correlation between CHADS_2_ score and the risk of CAC, with an incremental 23% increase in risk for each additional point on the score. However, no significant correlation was observed between severe coronary artery calcification and CHADS_2_ score, indicating potential differences in the underlying mechanisms of severe and non-severe calcified lesions in the coronary arteries. Therefore, further investigation with a larger sample size and additional features is warranted.

The pathological physiological process of vascular calcification is closely associated with hypertension and CHD. A meta-analysis involving 15,769 patients revealed that hypertensive individuals have a 1.71-fold higher risk of CAC compared to non-hypertensive individuals^15^, which aligns with our research findings. Furthermore, this study also suggests that hypertension can be considered as one of the predictive factors for severe calcification, potentially explaining the disparity in results between our study and theirs due to the inclusion of more significant variables. There exists a strong association between CHD and CAC, which is a concurrent phenomenon of severe progression in atherosclerosis^33^, and also serves as an essential predictor for CHD risk. This study once again elucidates the correlation between CHD and CAC, while exploring the impact of multiple variables on predicting both the occurrence and extent of vascular calcification. These findings provide valuable insights for clinical practitioners regarding whether additional evaluation of vascular calcification is warranted when managing patients with CHD.

The deposition of calcium phosphate, establishment of osteogenic signals, and disruption of calcium ion homeostasis are intricately associated with vascular calcification; however, the precise mechanism underlying this process remains incompletely elucidated^34–36^. Sustaining optimal levels of serum calcium consistently can mitigate the risk of calcification progression in dialysis patients^37^, underscoring the significant impact of serum calcium levels on CAC development. In this study, the normal range of serum calcium was found to be 2.11 to 2.52 mmol/L. Although no significant correlation was observed between serum calcium levels and coronary artery calcification, a clear negative correlation was found with severe coronary artery calcification lesions. This phenomenon may be attributed to the excessive transfer and deposition of serum calcium ions within blood vessels. However, limited research has been conducted on the correlation between low serum calcium levels and coronary artery calcification in non-chronic renal failure patients, necessitating further investigation.

Our model possesses several notable advantages. Firstly, we have innovatively incorporated CHADS_2_ score and carotid artery calcification as indicators for the prediction of CAC and sCAC, yielding commendable outcomes. Secondly, we have developed predictive nomograms for both CAC and sCAC, utilizing easily obtainable variables that hold strong practicality. Moreover, our model exhibits exceptional discrimination, calibration, and clinical applicability as validated by internal dataset.

Our study has several limitations. Firstly, given the dynamic nature of CACS, it is crucial to acknowledge potential time-varying confounding factors that cannot be disregarded, and causal relationships cannot be inferred. Secondly, this clinical study was conducted at a single center and lacks external validation, which may restrict its generalizability across different regions. Thirdly, our investigation focused solely on hospitalized patients with various reasons for admission, most of whom had multiple complications; moreover, stress reactions could also contribute to fluctuations in blood markers. More research is warranted to determine the applicability of this predictive nomogram in the general population. Additionally, individuals with confirmed CKD were initially excluded from our study; therefore, this predictive nomogram may not be suitable for such individuals. Despite these limitations, we have successfully developed two predictive nomograms that effectively estimate the risk of CAC and sCAC in middle-aged and elderly patients. We anticipate that these predictive nomograms can aid clinicians in early identification of elevated-risk individuals for CAC and sCAC while facilitating appropriate preventive measures or avoidance strategies.

## CONCLUSIONS

We have developed two nomograms that exhibit robust predictive performance for CAC and sCAC, enabling clinical physicians to make informed decisions regarding subsequent screening procedures.

## Non-standard Abbreviations and Acronyms

CAC: Coronary Artery Calcification
sCAC: Severe Coronary Artery Calcification
CACS: Coronary Artery Calcification Score
CT: Computerized Tomography
LASSO: Least Absolute Shrinkage and Selection Operator
CHD: Coronary Heart Disease
ROC: Receiver Operating Characteristic
AUC: Area Under the Receiver Operating Characteristic Curve
DCA: Decision Curve Analysis
CKD: Chronic Kidney Disease
RCS: Restricted Cubic Spline
CIC: Clinical Impact Curve
SD: Standard Deviation
IQR: Interquartile Range
OR: Odds Ratio
H-L: Hosmer-Lemeshow
CVD: Cardiovascular Disease
CTA: Computerized Tomographic Angiography
HDLC: Low-density Lipoprotein Cholesterol
TG: Triglyceride
LDL-C: Low-density Lipoprotein Cholesterol
HDL-C: High-density Lipoprotein Cholesterol
DM: Diabetes Mellitus
COPD: Chronic Obstructive Pulmonary Disease
ICVD: Ischemic Cerebrovascular Disease

## Declarations

### Ethics statement

The study was approved by the Ethics Review Committee of Taizhou Hospital of Traditional Chinese Medicine Ethics statement (Ethics Approval Number: 2024-033-01), and informed consent was waived. Before data analysis, all patient information was anonymized. The research conducted in this study adhered to the principles outlined in the Declaration of Helsinki.

### Consent for publication

Not applicable.

### Availability of data and materials

The datasets generated and/or analyzed during the current study are not publicly available due to hospital restrictions, but they can be obtained from the corresponding author on reasonable request.

### Competing Interests

The authors declare that they have no competing interests.

### Funding

The study was supported by grants from the Research Project on Elderly Health of Jiangsu Commission of Health (Jiangsu Province, China; LK2021058), and the Chinese Medicine Science and Technology Development Project of Taizhou Commission of Health (TZ202205).

### Authors’ contributions

Peng Xue analyzed the data, drafted the manuscript, and designed this study with Xiaohu Chen. Ling Lin and Yanshuang Zhuang were responsible for data collection under the supervision of Zhengting Deng. Peishan Li made substantial revisions to the manuscript. All authors read through and approved the final version. The corresponding author ensured that all listed authors met authorship.

## Acknowledgments

We would like to thank the staff from the Department of Geriatrics at Taizhou Hospital of Traditional Chinese Medicine for their assistance in data collection.

## Notes

### Competing Interest Statement

The authors have declared no competing interest.

## REFERENCES

1. Demer LL and Tintut Y. Vascular calcification: pathobiology of a multifaceted disease. Circulation. 2008;117:2938–48.

2. Greenland P, Blaha MJ, Budoff MJ, Erbel R and Watson KE. Coronary Calcium Score and Cardiovascular Risk. Journal of the American College of Cardiology. 2018;72:434–447.

3. Inoue K, Seeman TE, Horwich T, Budoff MJ and Watson KE. Heterogeneity in the Association Between the Presence of Coronary Artery Calcium and Cardiovascular Events: A Machine-Learning Approach in the MESA Study. Circulation. 2023;147:132–141.

4. Bourantas CV, Zhang YJ, Garg S, Iqbal J, Valgimigli M, Windecker S, Mohr FW, Silber S, Vries T, Onuma Y, Garcia-Garcia HM, Morel MA and Serruys PW. Prognostic implications of coronary calcification in patients with obstructive coronary artery disease treated by percutaneous coronary intervention: a patient-level pooled analysis of 7 contemporary stent trials. Heart (British Cardiac Society). 2014;100:1158–64.

5. Wasiak J, Law J, Watson P and Spinks A. Percutaneous transluminal rotational atherectomy for coronary artery disease. The Cochrane database of systematic reviews. 2012;12:Cd003334.

6. Lee MS, Gordin JS, Stone GW, Sharma SK, Saito S, Mahmud E, Chambers J, Généreux P and Shlofmitz R. Orbital and rotational atherectomy during percutaneous coronary intervention for coronary artery calcification. Catheterization and cardiovascular interventions : official journal of the Society for Cardiac Angiography & Interventions. 2018;92:61–67.

7. Blachutzik F, Meier S, Weissner M, Schlattner S, Gori T, Ullrich-Daub H, Gaede L, Achenbach S, Möllmann H, Chitic B, Aksoy A, Nickenig G, Weferling M, Dörr O, Boeder N, Bayer M, Elsässer A, Hamm C and Nef H. Comparison of Coronary Intravascular Lithotripsy and Rotational Atherectomy in the Modification of Severely Calcified Stenoses. The American journal of cardiology. 2023;197:93–100.

8. Newman AB, Naydeck BL, Sutton-Tyrrell K, Feldman A, Edmundowicz D and Kuller LH. Coronary artery calcification in older adults to age 99: prevalence and risk factors. Circulation. 2001;104:2679–84.

9. Liu Y, Fu S, Bai Y, Luo L and Ye P. Relationship between age, osteoporosis and coronary artery calcification detected by high-definition computerized tomography in Chinese elderly men. Archives of gerontology and geriatrics. 2018;79:8–12.

10. Feng W, Li Z, Guo W, Fan X, Zhou F, Zhang K, Ou C, Huang F and Chen M. Association Between Fasting Glucose Variability in Young Adulthood and the Progression of Coronary Artery Calcification in Middle Age. Diabetes care. 2020;43:2574–2580.

11. Bielak LF, Turner ST, Franklin SS, Sheedy PF, 2nd and Peyser PA. Age-dependent associations between blood pressure and coronary artery calcification in asymptomatic adults. Journal of hypertension. 2004;22:719–25.

12. Oshunbade AA, Kassahun-Yimer W, Valle KA, Hamid A, Kipchumba RK, Kamimura D, Clark D, 3rd, White WB, DeFilippis AP, Blaha MJ, Benjamin EJ, O’Brien EC, Mentz RJ, Rodriguez CJ, Fox ER, Butler J, Keith RJ, Bhatnagar A, Marie Robertson R, Correa A and Hall ME. Cigarette Smoking, Incident Coronary Heart Disease, and Coronary Artery Calcification in Black Adults: The Jackson Heart Study. Journal of the American Heart Association. 2021;10:e017320.

13. Lee MJ, Park JT, Chang TI, Joo YS, Yoo TH, Park SK, Chung W, Kim YS, Kim SW, Oh KH, Kang SW, Choi KH, Ahn C and Han SH. Smoking Cessation and Coronary Artery Calcification in CKD. Clinical journal of the American Society of Nephrology : CJASN. 2021;16:870–879.

14. Ono M, Takebe N, Oda T, Nakagawa R, Matsui M, Sasai T, Nagasawa K, Honma H, Kajiwara T, Taneichi H, Takahashi Y, Takahashi K and Satoh J. Association of coronary artery calcification with MDA-LDL-C/LDL-C and urinary 8-isoprostane in Japanese patients with type 2 diabetes. *Internal medicine (Tokyo*, Japan). 2014;53:391–6.

15. Nicoll R, Zhao Y, Ibrahimi P, Olivecrona G and Henein M. Diabetes and Hypertension Consistently Predict the Presence and Extent of Coronary Artery Calcification in Symptomatic Patients: A Systematic Review and Meta-Analysis. International journal of molecular sciences. 2016;17.

16. Nakamura S, Ishibashi-Ueda H, Niizuma S, Yoshihara F, Horio T and Kawano Y. Coronary calcification in patients with chronic kidney disease and coronary artery disease. Clinical journal of the American Society of Nephrology : CJASN. 2009;4:1892–900.

17. Hyun YY, Kim H, Oh KH, Ahn C, Park SK, Chae DW, Oh YK, Choi KH, Han SH, Kim YH and Lee KB. eGFR and coronary artery calcification in chronic kidney disease. European journal of clinical investigation. 2019;49:e13101.

18. Tang X, Qian H, Lu S, Huang H, Wang J, Li F, Bian A, Ye X, Yang G, Ma K, Xing C, Xu Y, Zeng M and Wang N. Predictive nomogram model for severe coronary artery calcification in end-stage kidney disease patients. Renal failure. 2024;46:2365393.

19. Park S, Hong M, Lee H, Cho NJ, Lee EY, Lee WY, Rhee EJ and Gil HW. New Model for Predicting the Presence of Coronary Artery Calcification. Journal of clinical medicine. 2021;10.

20. Wetscherek MTA, McNaughton E, Majcher V, Wetscherek A, Sadler TJ, Alsinbili A, Teh WH, Moore SD, Patel N, Smith WPW and Krishnan U. Incidental coronary artery calcification on non-gated CT thorax correlates with risk of cardiovascular events and death. European radiology. 2023;33:4723–4733.

21. Wang W, Wang H, Chen Q, Zhou Z, Wang R, Wang H, Zhang N, Chen Y, Sun Z and Xu L. Coronary artery calcium score quantification using a deep-learning algorithm. Clinical radiology. 2020;75:237.e11–237.e16.

22. Agatston AS, Janowitz WR, Hildner FJ, Zusmer NR, Viamonte M, Jr. and Detrano R. Quantification of coronary artery calcium using ultrafast computed tomography. Journal of the American College of Cardiology. 1990;15:827–32.

23. Gerke O, Lindholt JS, Abdo BH, Lambrechtsen J, Frost L, Steffensen FH, Karon M, Egstrup K, Urbonaviciene G, Busk M, Mickley H and Diederichsen ACP. Prevalence and extent of coronary artery calcification in the middle-aged and elderly population. European journal of preventive cardiology. 2022;28:2048–2055.

24. Mizuiri S, Nishizawa Y, Yamashita K, Mizuno K, Ishine M, Doi S, Masaki T and Shigemoto K. Coronary artery calcification score and common iliac artery calcification score in non-dialysis CKD patients. *Nephrology (Carlton*, Vic). 2018;23:837–845.

25. Homssi M, Vora A, Zhang C, Baradaran H, Kamel H and Gupta A. Association Between Spotty Calcification in Nonstenosing Extracranial Carotid Artery Plaque and Ipsilateral Ischemic Stroke. Journal of the American Heart Association. 2023;12:e028525.

26. Homssi M, Saha A, Delgado D, RoyChoudhury A, Thomas C, Lin M, Baradaran H, Kamel H and Gupta A. Extracranial Carotid Plaque Calcification and Cerebrovascular Ischemia: A Systematic Review and Meta-Analysis. Stroke. 2023;54:2621–2628.

27. van der Toorn JE, Bos D, Ikram MK, Verwoert GC, van der Lugt A, Ikram MA, Vernooij MW and Kavousi M. Carotid Plaque Composition and Prediction of Incident Atherosclerotic Cardiovascular Disease. Circulation Cardiovascular imaging. 2022;15:e013602.

28. Zhu G, Hom J, Li Y, Jiang B, Rodriguez F, Fleischmann D, Saloner D, Porcu M, Zhang Y, Saba L and Wintermark M. Carotid plaque imaging and the risk of atherosclerotic cardiovascular disease. Cardiovascular diagnosis and therapy. 2020;10:1048–1067.

29. Bos D, Arshi B, van den Bouwhuijsen QJA, Ikram MK, Selwaness M, Vernooij MW, Kavousi M and van der Lugt A. Atherosclerotic Carotid Plaque Composition and Incident Stroke and Coronary Events. Journal of the American College of Cardiology. 2021;77:1426–1435.

30. Stróżecki P, Serafin Z, Adamowicz A, Flisiński M, Włodarczyk Z and Manitius J. Coronary artery calcification and large artery stiffness in renal transplant recipients. Advances in medical sciences. 2015;60:240–5.

31. Ji C, Wu S, Shi J, Huang Z, Chen S, Wang G and Cui W. Baseline CHADS2 Score and Risk of Cardiovascular Events in the Population Without Atrial Fibrillation. The American journal of cardiology. 2020;129:30–35.

32. Caro-Codón J, Lip GYH, Rey JR, Iniesta AM, Rosillo SO, Castrejon-Castrejon S, Rodriguez-Sotelo L, Garcia-Veas JM, Marco I, Martinez LA, Martin-Polo L, Merino C, Martinez-Cossiani M, Buño A, Gonzalez-Valle L, Herrero A, Lopez-de-Sa E and Merino JL. Prediction of thromboembolic events and mortality by the CHADS2 and the CHA2DS2-VASc in COVID-19. Europace : European pacing, arrhythmias, and cardiac electrophysiology : journal of the working groups on cardiac pacing, arrhythmias, and cardiac cellular electrophysiology of the European Society of Cardiology. 2021;23:937–947.

33. Onnis C, Virmani R, Kawai K, Nardi V, Lerman A, Cademartiri F, Scicolone R, Boi A, Congiu T, Faa G, Libby P and Saba L. Coronary Artery Calcification: Current Concepts and Clinical Implications. Circulation. 2024;149:251–266.

34. Villa-Bellosta R. Vascular Calcification: Key Roles of Phosphate and Pyrophosphate. International journal of molecular sciences. 2021;22.

35. Sutton NR, Malhotra R, St Hilaire C, Aikawa E, Blumenthal RS, Gackenbach G, Goyal P, Johnson A, Nigwekar SU, Shanahan CM, Towler DA, Wolford BN and Chen Y. Molecular Mechanisms of Vascular Health: Insights From Vascular Aging and Calcification. Arteriosclerosis, thrombosis, and vascular biology. 2023;43:15–29.

36. Lee SJ, Lee IK and Jeon JH. Vascular Calcification-New Insights Into Its Mechanism. International journal of molecular sciences. 2020;21.

37. Zhang H, Li G, Yu X, Yang J, Jiang A, Cheng H, Fu J, Liang X, Liu J, Lou J, Wang M, Xing C, Zhang A, Zhang M, Xiao X, Yu C, Wang R, Wang L, Chen Y, Guan T, Peng A, Chen N, Hao C, Liu B, Wang S, Shen D, Jia Z and Liu Z. Progression of Vascular Calcification and Clinical Outcomes in Patients Receiving Maintenance Dialysis. JAMA network open. 2023;6:e2310909.

